# Dynamical and time series approach to understanding compartmental stock and flow models: a case study in malaria intervention models

**DOI:** 10.1101/2025.09.04.25335145

**Authors:** Eugene Tan, Mauricio van den Berg, Camilo Vargas, Peter W. Gething, Tasmin L. Symons

**Author notes:** **Author for correspondence:** Eugene Tan.

## Abstract

Insecticide treated nets (ITNs) represent one of the most cost-effective malaria control measures that is widely adopted today. The construction of mechanistic models to describe structural distribution, ownership and attrition of ITNs are crucial in order to quantify historical impact and burden, and perform predictive estimates of intervention impact. Compartmental stock and flow (SNF) models have been the traditional approach to modelling ITN inventories and have remained the most commonly employed approach owing to their simplicity and ease of implementation. However, insight into the mathematical justification for commonly adopted modelling decisions are sparse. The calibration of SNF to observed data is also challenging due as data across disparate sampling frequencies and sparsity need to be reconciled.

In this paper, we present a mathematical analysis of compartmental SNF models from both a time series analysis and dynamical systems approach to provide more insight on their dynamical behaviours. Using a reduced form of an SNF model, we show its equivalence to a linear time invariant system and demonstrate the criticality of attrition functions in the design SNF models. Additionally, we propose an iterative adapted expectation-maximisation (EM) algorithm to address SNF calibration challenges arising from disparate sampling frequencies alongside a list of required assumptions. Statistical analyses to verify the validity of these assumptions are presented. To the demonstrate its application, the subsequent EM method is applied to collected delivery, distribution and household survey data across 44 countries spanning 24 years to provide robust and statistically rigorous estimates of net distribution and volumes. Results for numerical convergence and uniqueness of outputs are also given.

## 1. Introduction

Compartmental models consist of a large class of approaches that are commonly employed in the study of biology and epidemiology. The latter application which this paper will focus on, particularly in the study of infectious disease spread and dynamics, has become one of the quintessential applications of compartmental approach since the seminal paper written by Kermack & McKendrick [1]. Since then, the compartmental modelling framework has seen numerous extensions to account for a myriad of different disease dynamics such as SIRS, SIRD and SEIR in the cases of pre-exposure, disease mortality, immunity [2–4]. The framework has also been used to model multiple competing strains [5,6], epidemics on social networks [7,8] and simplicial contagion [9,10].

The prominence of compartmental models can be partially attributed to the flexibility and interpretability of the compartmental approach. However, it is for a similar reason that such methods can be mathematically difficult to analyse owing to the intractabilty of solutions despite the governing equations having relatively simple forms. Whilst it is possible to write down solution approximations for some simple cases, most applications utilise numerical simulations to rapidly produce solution trajectories [11]. A bigger challenge instead is deciding on model choices, particularly when trying to describe a novel context where the standard mathematical formulations (e.g. exponential growth and decay, and product interaction terms) may not be appropriate. This challenge is further compounded if there is a requirement to calibrate model parameters against observed data.

An area where this problem arises is in the particular context of malaria intervention models for insecticide treated bed nets (ITNs) where the compartmental approach is also known as a “stock-and-flow” (SNF) model. Despite its downstream impact and relevance in the study and policy design of interventions [12–14], the design of ITN models have traditionally employed ad hoc approaches which have not been formally mathematically justified in the literature. In this paper, we analyse a compartmental SNF model based on those presented by Bertozzi-Villa et al. using a dynamical systems and time series analysis approach and provide some mathematical justifications on the design choices for one of its key components: the attrition function. Furthermore, we also present a statistically conservative approach to improving the fit and temporal resolution of SNF models when calibrating against observation data containing differing sampling resolutions and propose an adapted expectation-maximisation (EM) algorithm for model calibration.

This paper is structured as follows. We first provide in Section 2 a historical overview of compartmental SNF models in the malaria ITN context is provided accompanied with a mathematical description of the full SNF model. In Section 2(c), we explore a mathematical reduction of the full SNF model. This reduction is employed in Section 3 to provide insight and mathematical justification on the importance of attrition functions in SNF model design. Section 4 discusses the problem of disparate sampling resolutions in calibration data and proposes a solution to this problem using temporal disaggregation. The proposed approach is applied in Section 5 followed by a discussion on the convergence behaviour and impact of different parameters on model fit and dynamics.

## 2. Malaria ITN Models

### (a) ITN Coverage Models

Since the first major efforts for malaria control in the mid 2000s, there has been enormous progress in the fight against malaria resulting in consistent decreases in childhood mortality and case incidence globally [14–16]. Among the three major forms of interventions, artemisinin recombinant therapy (ACT), indoor residual spraying (IRS) and insecticide treated nets (ITNs), much of the progress in reducing the malaria disease burden has been attributed to ITNs [13]. However, a slowdown of progress from the year 2018 coupled with large changes in the economic and epidemiological landscape has prompted increased interest to revisit existing standardised model for malaria interventions [17–19].

The main aim of ITN coverage models is to track, estimate and predict the status of ITNs within communities to gain insight on their respective levels of access, usage and protection. These models are highly useful in the design of policies as they allow tracking of current progress, and in the case of predictive models, a way to simulate and forecast the impact of different intervention programme strategies [20,21]. Futhermore, ITN coverage models form the basis for estimating historical impact and burden estimates of malaria and relevant interventions [14,15].

Several ITN coverage models have been proposed and deployed since the rollout of national ITN distribution campaigns for malaria intervention. The earliest of these is the NetCALC model first presented by Killian et al. [22]. The NetCALC model is a publicly available spreadsheet-based model primarily used assist with the study of net procurement and durability. However, this model is not amenable to historical analysis of existing data, or more advanced statistical methods.

One of the first attempts at mathematically modelling and estimating ITN crop from surveys using historical data was proposed by Flaxman et al. in 2010 [23]. They proposed the usage of a compartmental model, described as a “stock-and-flow” approach, to track and simulate the movement of nets across various life stages (national inventory, in community, and discarded) within a given country over time. The Flaxman model also standardised the usage of delivery data alongside reported distributions in order to provide more robust estimates of actual net distributions into the community. However, fitting the model required various constraints and limitations such as aggregating data into a coarse annual time step, and assuming that all nets were discarded after 3 years of life. Furthermore, the model defined ITN coverage as “household ownership of at least one ITN”, which is no longer used. Instead, current analyses define coverage in terms of ITN access, defined as “one net available between two people”.

An extension of the Flaxman model was presented by Bhatt et al. in 2015 [13]. The Bhatt model contained improvements such as increasing the temporal resolution of the model to quarterly times steps to better match the monthly resolution of household surveys. Furthermore, they also introduced a proper loss function in the form of a two parameter compact exponential decay to describe net attrition, as well as the calculation of five different ITN metrics: household ownership, household access, population access, population use and ownership gap. Bhatt et al. model presented an initial attempt to account for missing net distribution data through the usage of spline interpolations. Whilst the above improvements provided flexibility and a degree of interpretability to the model, insight on the identifiability of model parameters was unclear.

Leveraging on the work done by Bhatt et al., Bertozzi-Villa et al [12] iterated on the model to allow for rudimentary imputation of missing data, and simplification of the loss function to provide better interpretability and identifiability whilst retaining a majority of the structure of the Bhatt model. The Bertozzi-Villa (BV) model was also extended to include location data from surveys for the purposes of geospatial disaggregation. This was used effectively to construct spatiotemporal regression models of net access and use across the African continent, and produce high resolution maps of net coverage across the time period of 2000-2020. Additionally, the BV model was the first attempt of producing a publicly available codebase, allowing for better review and reproducibility.

Currently, the BV model remains the gold-standard approach for ITN coverage modelling and has been used in numerous policy and downstream modelling exercises [24]. In its essence however, main driving mechanism of the BV model has its roots in the underlying SNF model, which has remained relatively unchanged since the form proposed by Bhatt et al.

### (b) Bhatt-BV Stock-and-Flow (SNF) Model

The stock-and-flow model used by Bhatt et al. and Bertozzi-Villa et al., hereby abbreviated as SNF for brevity, aims to simulate the flow of ITN through a system (i.e. country) and describes two main model components. Firstly, the SNF model tries to estimate the effective number of nets distributed by a country within a given period of time. In more sophisticated formulations, these can include a rudimentary stockpiling mechanism to account for errors in reported distribution data and a imputation of both net delivery and distribution data. Once net distribution volumes have been established, the second component models the natural attrition of nets within the community to estimate the volume of actively utilised nets. We clarify that the usage of the term “attrition” does not necessarily refer to the failure of nets due to reliability or wear, but instead it refers to the general the disappearance of nets from the community.

The general ITN SNF model can be written explicitly as a set of non-autonomous difference equations that describe the net distribution and attrition separately. The net distribution portion that reconciles distribution and delivery using a gating mechanism are given by the following equations,

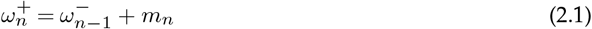

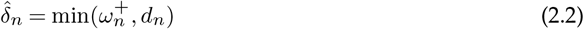

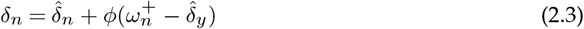

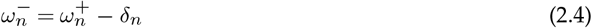

subject to the initial condition,

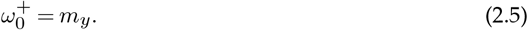

The variables in the above equation are defined as follows:

- *m*_*n*_ : Reported number of ITNs delivered at the start of year *n*
- *d*_*n*_ : Reported number of ITNs distributed at the start of year *n*
- 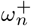 : Number of ITNs in the national stock at the start of year *n*
- 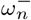: Number of ITNs in the national stock at the end of year *n*
- 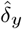: Number of ITNs distributed excluding additional redistribution of current excess net stock
- *δ*_*y*_ : Number of actual ITNs distributed during the entirety of year *n*

In this form, it is assumed that reported distributions are not completely accurate and true distributions cannot be greater than the amount of available nets in inventory as indicated by reported deliveries. Should there be excess nets available, a portion *ϕ* is used to supplement reported net distributions to provide a final distribution time series *δ*_*y*_. which is subsequently used as the driving signal for the attrition component of the model. It is important to note that part of the flexbility in SNF lies in the choice of gating mechanism that govern the output driving signal.

Traditionally, the attrition component of SNF models track each cohort of distributed nets separately,

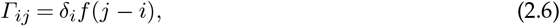

where *Γ*_*ij*_ is the number of surviving nets in the community at time *j* that were distributed at time *j*, and *f*(*t*) is the attrition survival function. In the simplest case, the choice of a memoryless attrition function such *f*(*t*) = *e*^−*kt*^ reduces the SNF model into a difference equation,

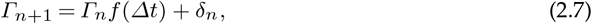

where Δ*t* is the size of time interval between the *n*^*th*^ and (*n* + 1)^*th*^ time step. However, this choice of attrition function while mathematically convenient does not reflect observed attrition relationships [22]. In practice, various sigmoidal shaped functions may be used the choice of which is a key design parameter of the SNF model, which we argue should be well justified. A discussion on the choice of attrition functions is provided in Section 3.

### (c) Reduced SNF Model

To better understand the governing dynamics of the SNF model, we consider two different reduced forms of the SNF model. In both cases, we assume that the effects of net redistribution due to the parameter *ϕ* is temporally local and small in magnitude compared to the underlying distribution time series *δ*_*y*_. Under this assumption, the dynamics of the SNF model is restricted to that of the attrition component.

The first is a dynamical systems approach where we first represent the SNF difference equation as a generalised discrete map,

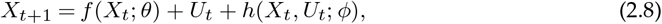

where *X*_*t*_ is the net stock in month *t, f* is the attrition survival function parameterised by *θ, h* are the dynamics associated with redistribution of excess net stock, and *U*_*t*_ = *g*(*δ*_*t*_) is a driving signal heavily correlated with the effective net distribution expressed as a transformation *g* of the reported net distribution *δ*_*t*_. Under the assumption the net deliveries and net distributions do not significantly differ (i.e. *ϕ* effects are negligible), the simplification of ||*h*(*X*_*t*_)|| ≈ 0 yields the following form,

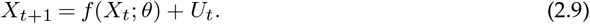

Assuming the decrease in net crop within a given time step Δ*t* is small relative to overall net crop,

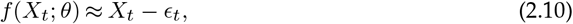

we get the simplified non-autonomous discrete map,

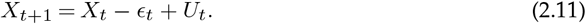

Taking the limit as Δ*t* → 0, a continuous analogue can be written to express the SNF as a non-autonomous ODE dynamical system,

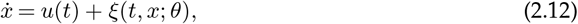

where the net distribution time series *u*(*t*) is the driving signal, and *ξ*(*t, x*; *θ*) are the net attrition dynamics. We note that this form is not directly useful for simulation. Instead, it allows for a potential explanation on the observed convergence behaviour when calibrating SNF models to data as discussed in Section 5(b).

An alternative approach to reducing the SNF model afforded by assuming net redistribution effects are negligible is to represent the SNF as a linear time invariant system. In this approach, the output time series of net crop *Γ*(*t*) can be represented as the forward convolution of an input distribution time series *δ*(*t*) and an impulse response described by the attrition function *f*(*t*),

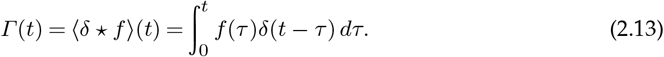

This representation provides a computationally simple way to simulate an approximation of the full SNF model and is useful isolating the effects of various parameters in the attrition model.

## 3. Attrition Functions

### (a) Functional Forms

The choice of attrition function *f*(*t*) greatly influences the output dynamics of SNF models as is evident in both reduced forms described in Section 2(c). Apart from the basic condition that *f*(*t*) must be monotonically decreasing, there is little restriction on its mathematical form. Excluding the overly simple exponential decay function, several other mathematical forms have been proposed [14], most of which are sigmoidal in shape. Here we consider two options of attrition function and provide justification for their usage when designing SNF models.

The first option is the Weibull distribution, a flexible flexible two parameter model commonly employed in survival analysis and reliability engineering to model the failure time [25]. The associated survival function is given by,

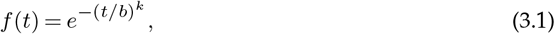

where *b* and *k* are the location and shape parameter respectively. The function has full support over the reals and is a generalisation of the exponential distribution.

Weibull models are able to capture three main modes of failure depending on the value of *k*. For *k <* 1, *f* ^*′*^(*t*) is strictly decreasing and younger components have a higher risk of failure. This is used to describe cases of infant mortality where failure risk decreases with age. For *k* = 1, the model reduces to exponential failure corresponding to a constant rate of failure. For *k >* 1, ℒ (*t*) is sigmoidal with an inflection point corresponding to a time of high failure risk.

An alternative to the Weibull model is the smooth and compact exponential loss function first proposed by Killian et al. [22] and subsequently used in the development of NetCALC and models by Bhatt et al. and Bertozzi-Vila et al [12,13]. They proposed the usage of a two parameter smooth and compact exponential loss function to model describe the natural attrition of nets with validation done against observed empirical data. This function is given by,

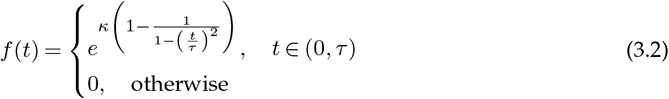

where *τ* and *κ* are model parameters. The time scale parameter *τ* directly restricts the support of *f*(*t*) and *κ* controls the decay shape. The restricted domain due to *τ* also allows the model to describe sudden onset failure modes when *κ* is small (see Figure 1). Both functions are asymmetric about its inflection point, with the compact exponential having a slightly greater degree of asymmetry particularly when *κ <* 1. However, this can also be achieved in the Weibull to a certain degree by scaling the function parameters.

**Figure 1.**
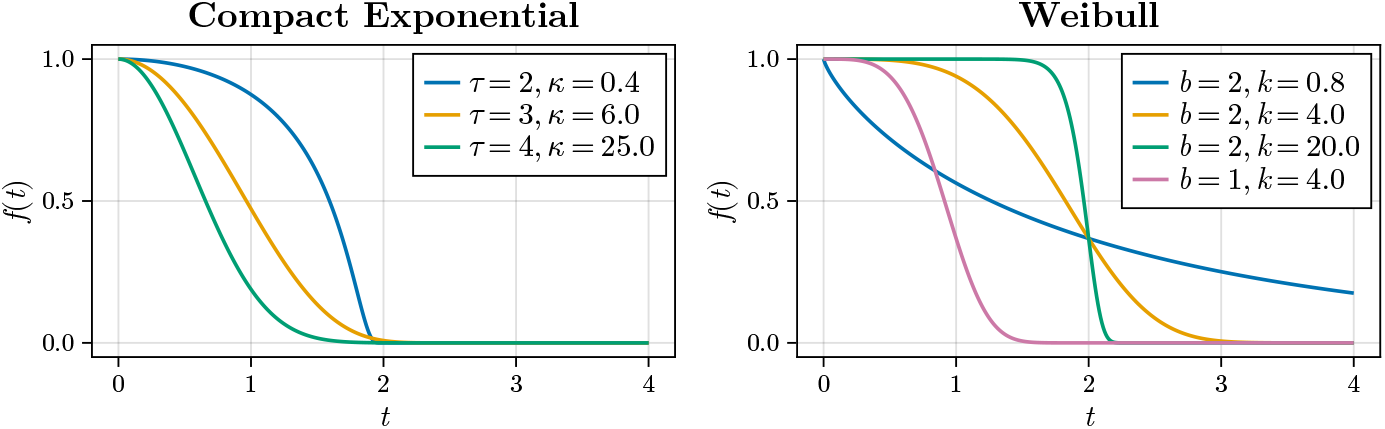
Characteristic shapes of the compact exponential and Weibull loss functions for various parameter values

Previous models of ITN coverage have used variations of the smooth compact exponential. The Bhatt model utilised the loss function in its full two parameter form when performing model fit. In later analyses, the BV model suggested that the two parameter form might affect parameter identifiability and utilised a simplified one parameter version with arbitrary *κ* = 20 held constant. The smooth compact exponential has also been used to represent net attrition generalised malaria simulation models such as OpenMalaria [26,27].

Apart from the requirement of a sigmoidal shape for net attrition, motivations for choosing the smooth compact exponential over other loss functions are not clear. Bhatt et al. adopted an empirical approach to choosing the loss function, using Bayesian model selection to select between smooth compact loss, Hill, and Weibull functional forms. The smooth compact loss function was found to produce lower Deviance Information Criteria (DIC) when assessed against their data, and was thus chosen [13]. In an effort to provide more insight on the choice of attrition parameter, we explore the effects of both candidate attrition functions’ parameters on SNF outputs and provide a brief discussion on the potential merits of each choice.

### (b) Quantifying attrition parameter effects on dynamics

We use the linear time invariant reduced form of the SNF model to compare the difference in output dynamics for both the Weibull and compact attrition curves. Two basic forms of distribution time series are tested: (1) temporally uniform distributions *δ*(*t*) = *k* representative of the routine distributions throughout a given year, and (2) seasonal annual distributions representative of policy strategies that try to preempt heavy case load seasons. The distribution time series for the latter is represented by a the periodic function,

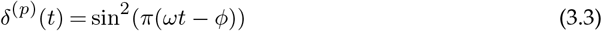

where the period is modulated by *ω* (number of cycles per time unit). Distribution quantities for both cases are normalised and dimensionless. To ensure that both cases (uniform *δ*^(*u*)^(*t*) and periodic *δ*^(*p*)^(t)) correspond to the same total number of input nets, distribution time series are scaled such that the following condition holds,

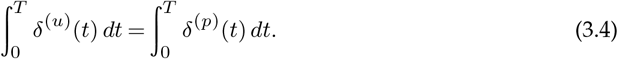

Several realisations for both the Weibull and compact exponential cases are shown in Figures 2 and 3 across a range of chosen parameters.

**Figure 2.**
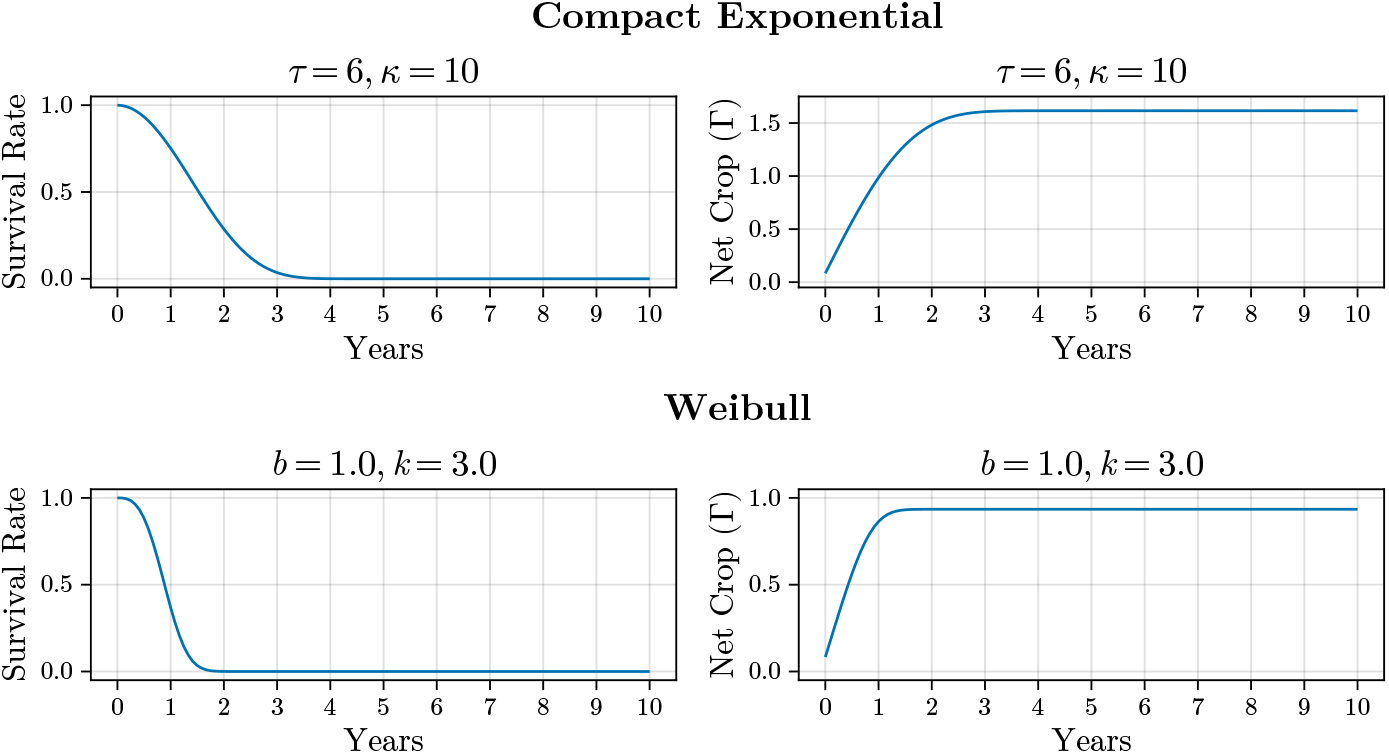
Simulated net crop trajectories with uniform annual distribution.

**Figure 3.**
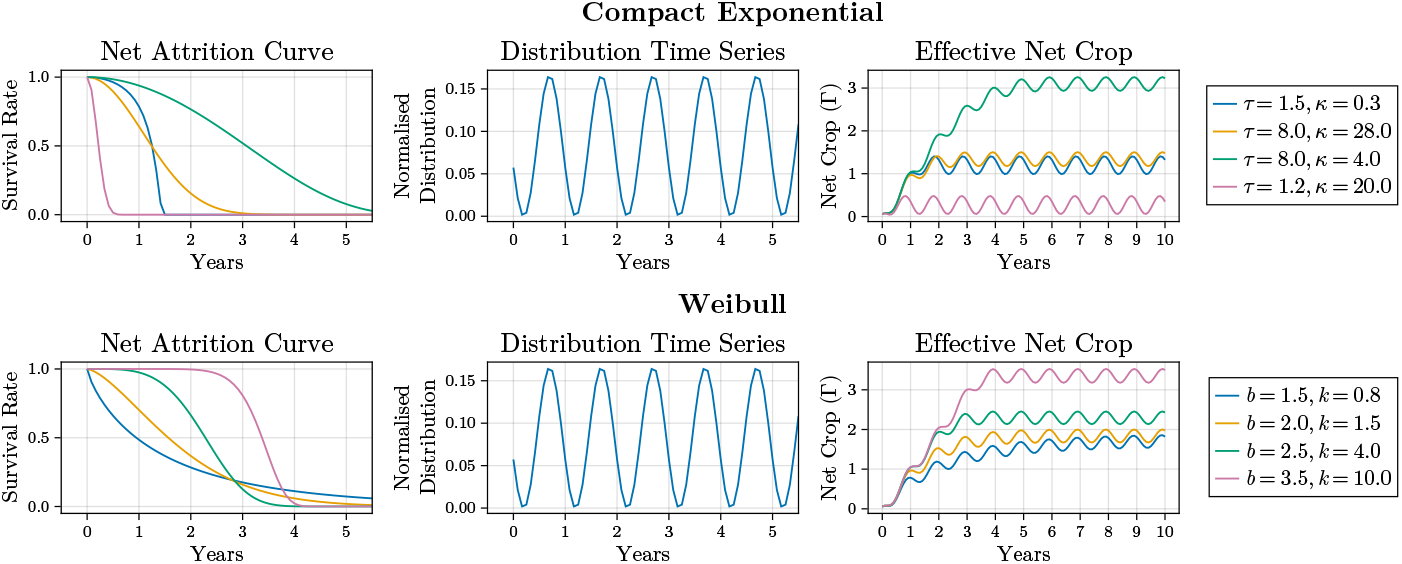
Simulated net crop trajectories with periodic annual distributions (*ω* = 1) for various attrition parameter combinations.

To better understand the effect of parameters in attrition function, we define two measures to summarise the outputs: (1) the steady state mean 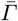, and (2) signal standard deviation *σ*_*Γ*_. For any monotonically decreasing function, the system will settle at a given steady state value 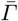. In the case of a periodic input, the output signal will oscillate about a mean of 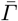. In the context of ITNs, this corresponds to the expected level of net crop availability in the community relative to a chosen distribution strategy. The second measure, signal standard deviation *σ*_*Γ*_, is only relevant for the oscillatory case and is calculated as the standard deviation of the output time series when the system has entered the steady oscillation regime. In this case, the standard deviation describes the expected amount of volatility in net availability, where large standard deviations are undesirable as it can result in periods of highly compromised ITN coverage in the community.

Parameter ranges of *κ, k* ∈ (0, 54.6) and *τ, b* ∈ (0, 5) are examined for the compact exponential and Weibull cases. These parameter ranges were chosen to cover the variety of characteristically different decay shapes afforded by each attrition function with sufficient flexibility. In all cases, simulations were run for a length of 30 years to ensure that steady state (or oscillation in the case of periodic inputs) is achieved. Phase plots of the uniform distribution case is shown in Figures 4, and for the periodic distribution case in Figures 5 and 6.

**Figure 4.**
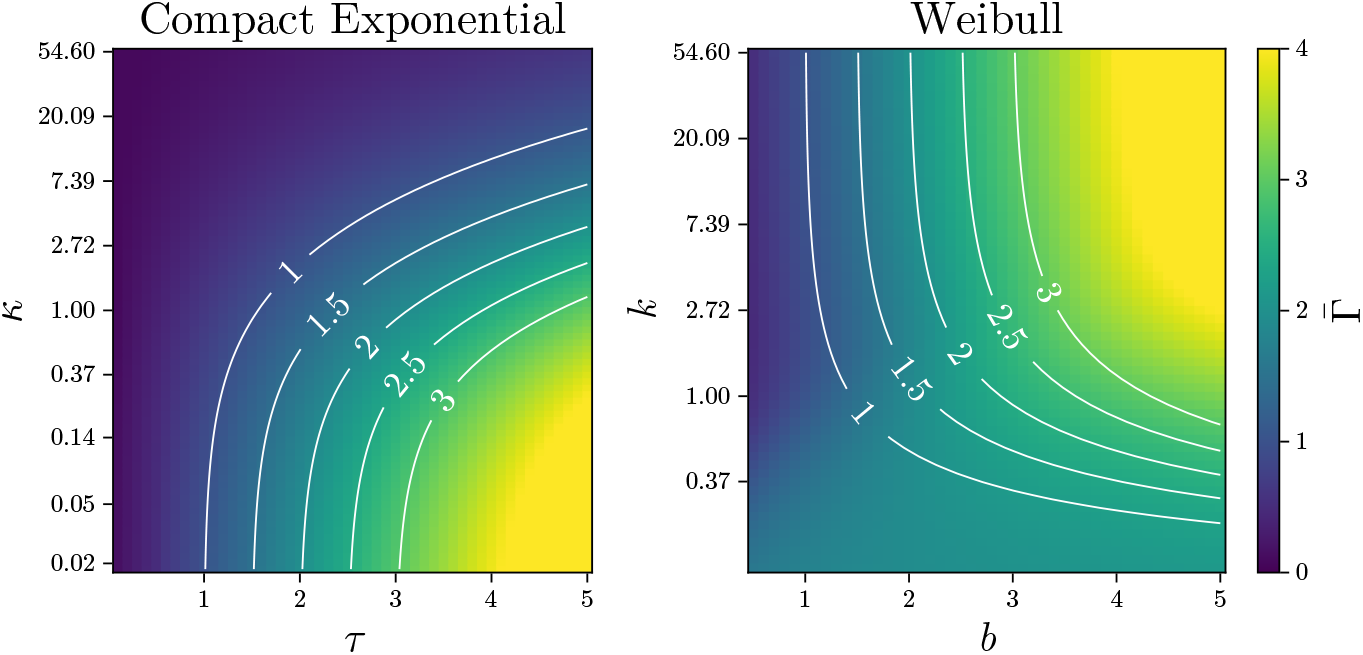
Phase plots of steady state net crop 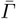 at various attrition parameter values. Contours indicate lines of constant attrition half life in years.

**Figure 5.**
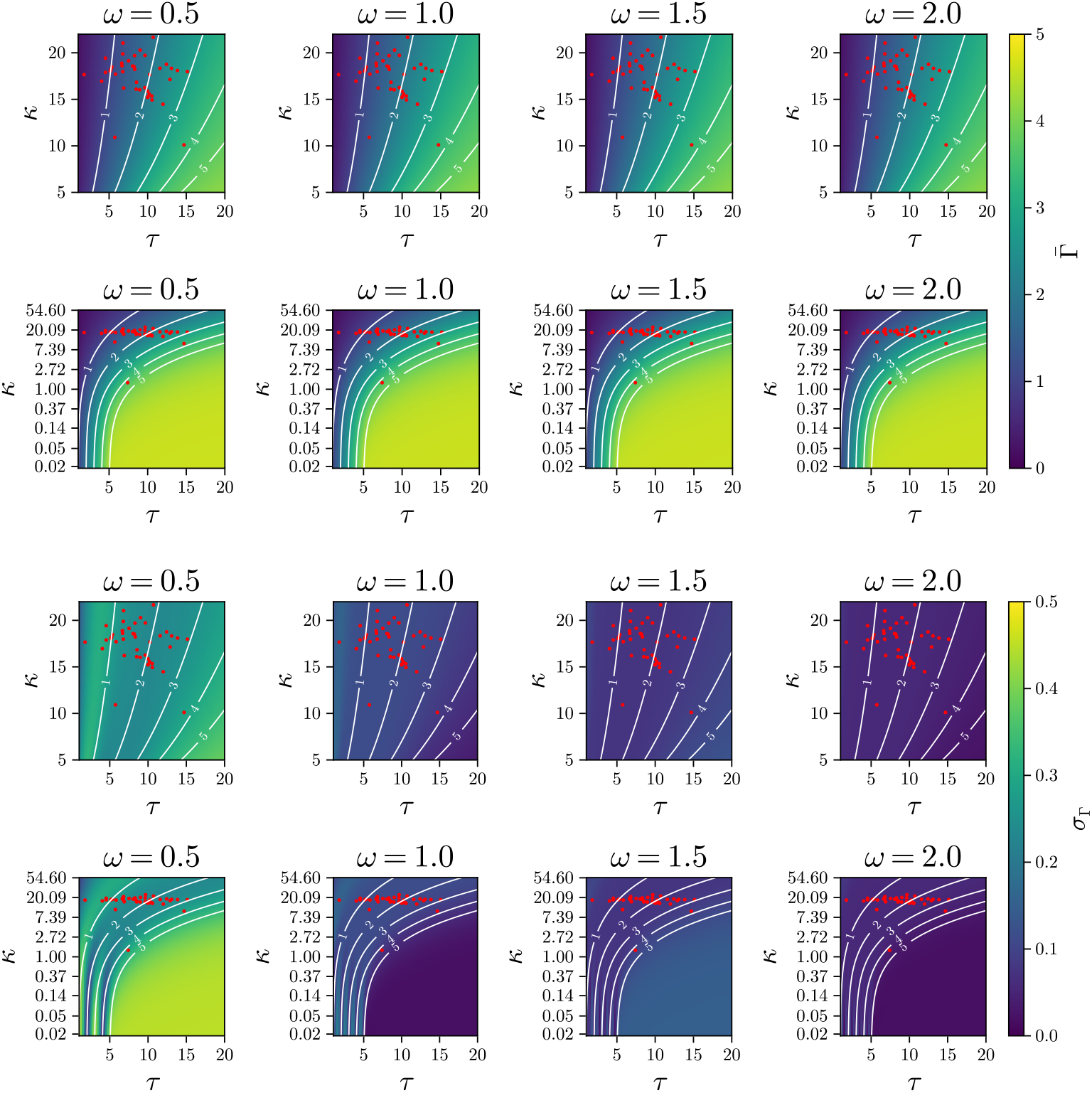
Phase plots for compact exponential attrition of 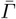 and *σ*_*Γ*_ at various attrition parameter values with periodic distribution *ω* = (0.5, 1, 1.5, 2). Contours indicate lines of constant attrition half life in years. Red dots are posterior estimates of SNF model attrition parameters when trained on real household survey data. Label scales for *κ* are given in linear and log scales for clarity.

**Figure 6.**
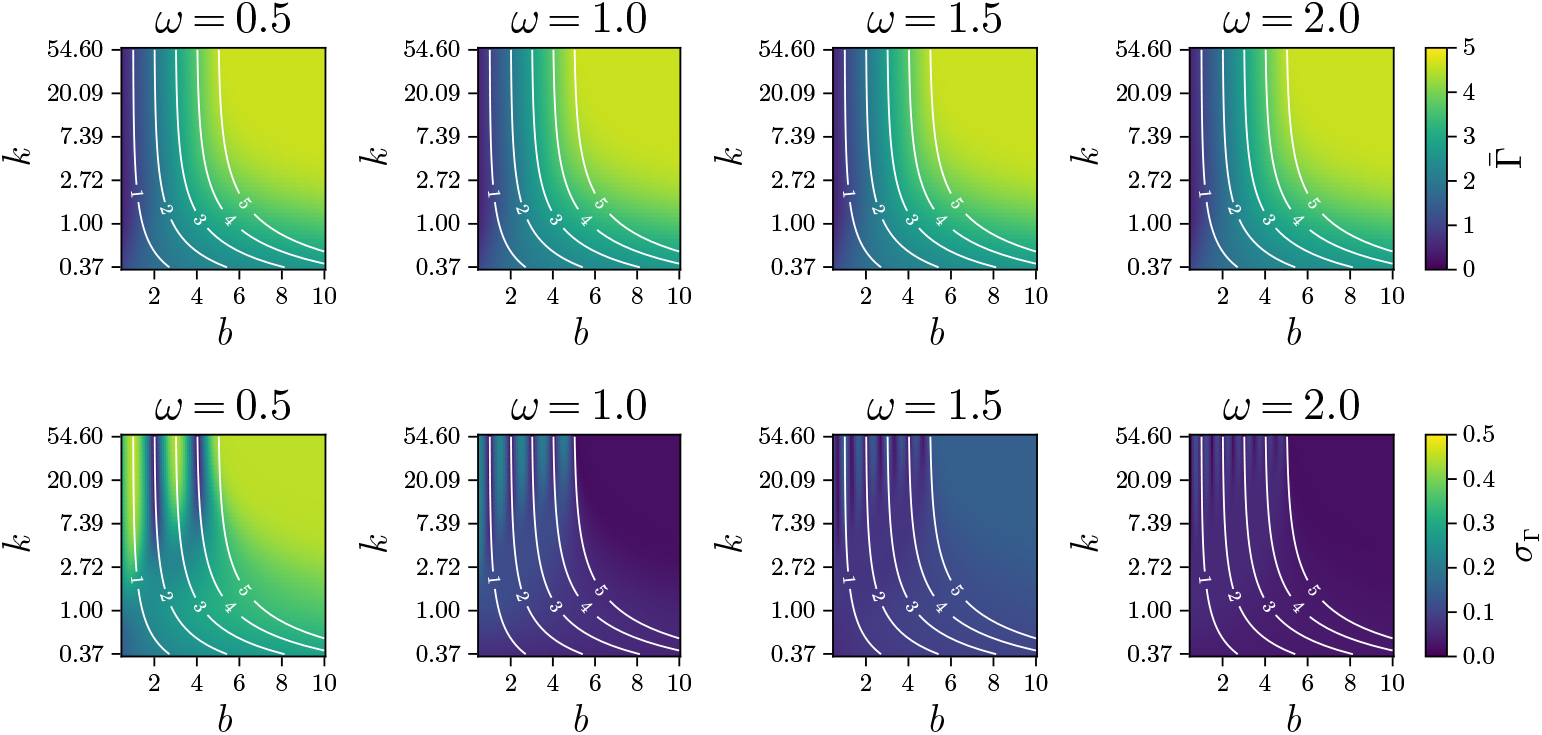
Phase plots for Weibull attrition of 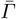 and *σ*_*Γ*_ at various attrition parameter values with periodic distribution *ω* = (0.5, 1, 1.5, 2). Contours indicate lines of constant attrition half life in years.

In all cases tested, it is unsurprising that attrition half-life is strongly correlated with the steady state mean 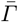 (see Figures 4, 5 and 6). However, both bairs of parameters in the compact exponential and Weibull attrition functions have a non-negligible effect on the 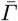. Generally, attrition curves with more prolonged high survival rates result in a higher steady state mean.

More interesting is the joint importance of parameters on *σ*_*Γ*_ in the periodic distribution case where resonance bands appear in at critical parameter ranges (see Figures 5 and 6). The attrition parameters in this regime mostly correspond to attrition curves that experience a sharp decline in survival at a critical time period. The time scales at which this occurs appears to interact with the natural frequencies of the attrition function producing alternating bands of high and low volatility. While these bands are interesting, the attrition curves do not correspond to typical observed attrition behaviour of ITNs, which exhibit a more gradual decay. However, the presence of these bands do not entirely disappear and there is still a non-negligible joint effect of parameters on volatility when considering longer periodicities in typical parameter ranges inferred from househould data (see red points for *ω* = 0.5 case in Figure 5).

We note that the qualitative behaviours of both the Weibull and compact exponential attrition functions are similar. More importantly, the joint importance of both parameters on 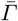 and *σ*_*Γ*_ is apparent in both cases. This observation is supported by the relatively wide variation in posterior parameter values when fitting the a full SNF model with compact exponentials on observed survey data (see Figure 5). These observations suggest that rather than the choice of mathematical form, it is the flexibility afforded by the number of attrition parameters that have a bigger impact on the resulting dynamics. Therefore, instead of the one-parameter simplified form adopted by Bertozzi-Villa et al. and Flaxman et al., a minimum of two parameters that allows control over the time scale and shape of the attrition function should be used to ensure sufficient flexibility when performing model calibration. The analysis in the rest of the paper will focus on the case where the attrition function is a compact exponential, but note that findings may be generalised to other attrition functions.

## 4. Temporal Disaggregation and Mismatched Resolutions

### (a) Resolution Mismatch and Temporal Disaggregation

Since Flaxman et al., the calibration of SNF models for ITNs have traditionally involved the reconciliation of a combination of data from various sources, namely, manufacturer ITN delivery to governments, national household ITN distributions, and household level surveys of ownership and use. This multimodal approach allows for realistic and reasonable constraints to be applied when calibrating attrition parameters. However, a challenge arises when differing data sources contain inherently different temporal resolution, often by nature of their method of data collection.

In the case of ITN tracking, the temporal resolution mismatch arises between the distribution and household surveys of net ownership and use. Delivery and distribution data is typically aggregated into regular annual intervals. In contrast, the time and economic cost of running household level surveys often results in observation data that is episodic and sparse with each survey consisting of a sequence of consecutive observations with high temporal resolution (monthly). Previous attempts to reconcile this problem typically involve aggregating or disaggregating data sources into a common intermediate resolution. For ITNs, this involves aggregating surveys into a single summary observation associated to one point in time. The opposite is done for delivery and distribution data where annual values are disaggregated uniformly into quarter-year periods with some small amount of randomness included for variation between quarters.

This approach of reducing datasets into an intermediate resolution poses several challenges unique to the ITN context. Firstly, the aggregation of survey data into a single time point discard much of the temporal detail inherent in the survey. Given survey time periods (4-7 months) and net half-lives (12-24) months have relatively comparable time scales, aggregation of survey data into a single observation ignores the simultaneous net attrition that occurs during the conducting of the survey the effect of which may be non-negligible. Secondly, while the uniform disaggregation of distribution data is straightforward, it fails to account for potential seasonal distribution strategies or the non-standard timing of mass campaigns that occur within a very short period.

Given that the lowest temporal resolution (annual) is too coarse to adequately describe the attrition dynamics of ITNs, an alternative solution to reconcile mismatched resolutions is to disaggregate all data sources into the highest temporal resolution amongst all data sources. For ITN models, this task involves the inference of monthly ITN distributions to produce a set of data that is temporally aligned and allows for more confidence in the estimation of net attrition parameters. This can be done by introducing a set of model parameters *p*_*n,m*_ representing the proportion of reported distributed nets in year *n* that were distributed in month *m* of that year. This is given by,

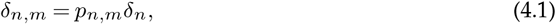

subject to the conservation condition,

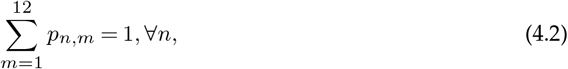

where *δ*_*n*_ is the number of nets distributed during year *n*. For convenience, the time index of net distributions can be rescaled to produce a monthly distribution time series *δ*_*t*_ where *t* = 12*n* + *m*.

Rather than the uniform disaggregation approach employed in existing BV and Bhatt models, we argue that disaggregation ratios within each year should be informed by observed data. However, high resolution disaggregation can result in a large explosion of model parameters that while all having only temporally local effects, requires calibration. In the subsequent sections we describe the conditions required for the validity of such a disaggregation in the ITN modelling context, and present a modified expectation-maximisation (EM) approach to perform such a fit.

### (b) Temporal Homogeneity Considerations

A problem that is unique to the study of SNF models for ITN is the validity of disaggregating household surveys at a monthly resolution. The construction of household survey samples are typically designed to be nationally representative and accounts for factors such as household size and rurality [28]. Therefore, aggregate metrics calculated over the entirety of the survey should be reflective of a statistic averaged across the entire country’s demographic. However, the scale of household survey operations often require multiple months to complete, during which substantial net attrition can occur. This can affect the true value of the calculated statistic.

Because surveys are only guaranteed to be nationally representative when considered in its entirety, a given month’s aggregate value may not be nationally representative depending on how the survey execution strategy (e.g. if household sampling was procedurally conducted by state). This can cause over-representation of certain regions during specific months of a survey and can distort monthly aggregate estimates. Therefore, any temporal disaggregation exercise based on survey entries requires testing on whether subsets of the survey entries remain nationally representative.

One way to test the validity of monthly aggregates is to check that the spatial sampling distribution for each month within a given survey campaign is both consistent across each month (temporal homogeneity), and also matches the sampling distribution when the survey is taken in its entirety. We note that such an approach does not guarantee sampling homogeneity when comparing across different social demographics within a given region (e.g. wealth and household size). However, given the correlations between geographical factors such as rurality, and many other social metrics, we argue that checking for temporal homogeneity should also provide a degree of implicit validity on the monthly aggregates across said social factors.

For each set of survey samples *S*, the geographical coordinates of each sample 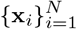 are used as centres for Gaussian kernels *Φ* to produce the following spatial distribution,

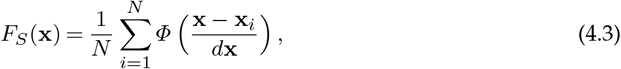

where *d***x** is a chosen bandwidth for kernel. Differences between two spatial distributions 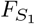 and 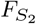 can be calculated using the Jensen-Shannon divergence (JSD) [29],

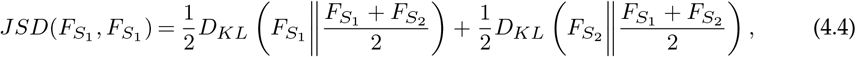

where *D*_*KL*_(*F*∥*G*) is the KL-divergence between any two distributions and is calculated as,

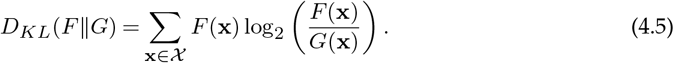

Given any pair of distributions on some domain 𝒳, the 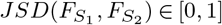 where a value of 0 corresponds to equality between distributions. Conversely, a value of 1 corresponds both distributions being fully disjoint.

To assess the temporal homogeneity of each survey, let *S*_*survey*_ correspond to the full collection of survey entries, and *S*_*i*_ be the subset of the survey entries that occur in the *i*^*th*^ month the survey (i.e. *S*_*survey*_ = ⋃_*i*_ *S*_*i*_). For each month in a given survey, we calculate the score *H* = 1 − *JSD*(*S*_*i*_, *S*_*survey*_) with respect to a spatial resolution of *d***x** equal to 1*/*50 of the latitude and longitude range in the dataset. Higher scores of *H* represent strong agreement between a given month’s sampling distribution and that of the entire survey. A comparison of the spatial distribution and *H* scores for a 2013 survey in Nigeria is provided in Figure 7. A summary of homogeneity scores for a collection of surveys in 2000-2023 across 44 analysed countries are shown in Figure 8.

**Figure 7.**
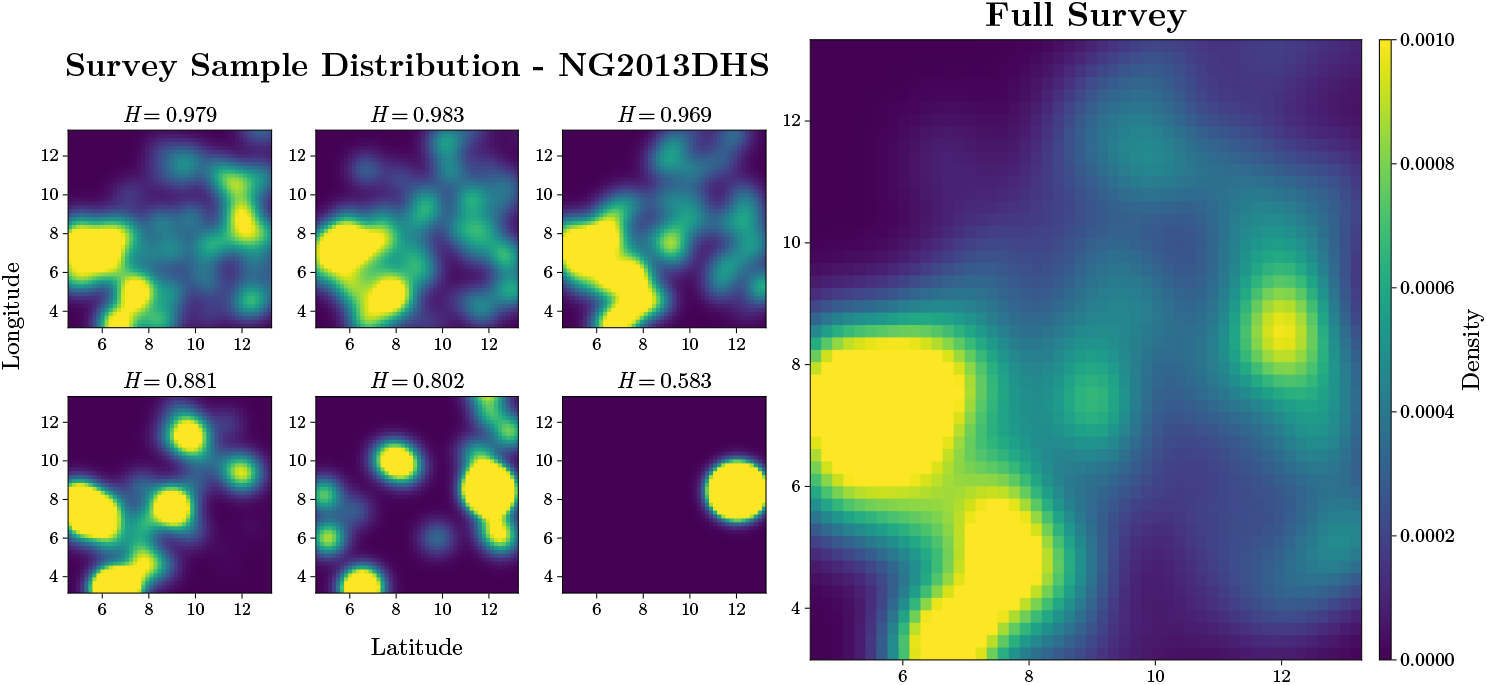
Monthly homogeneity scores and spatial distribution of household samples compared against full survey for the 2013 DHS survey in Nigeria.

**Figure 8.**
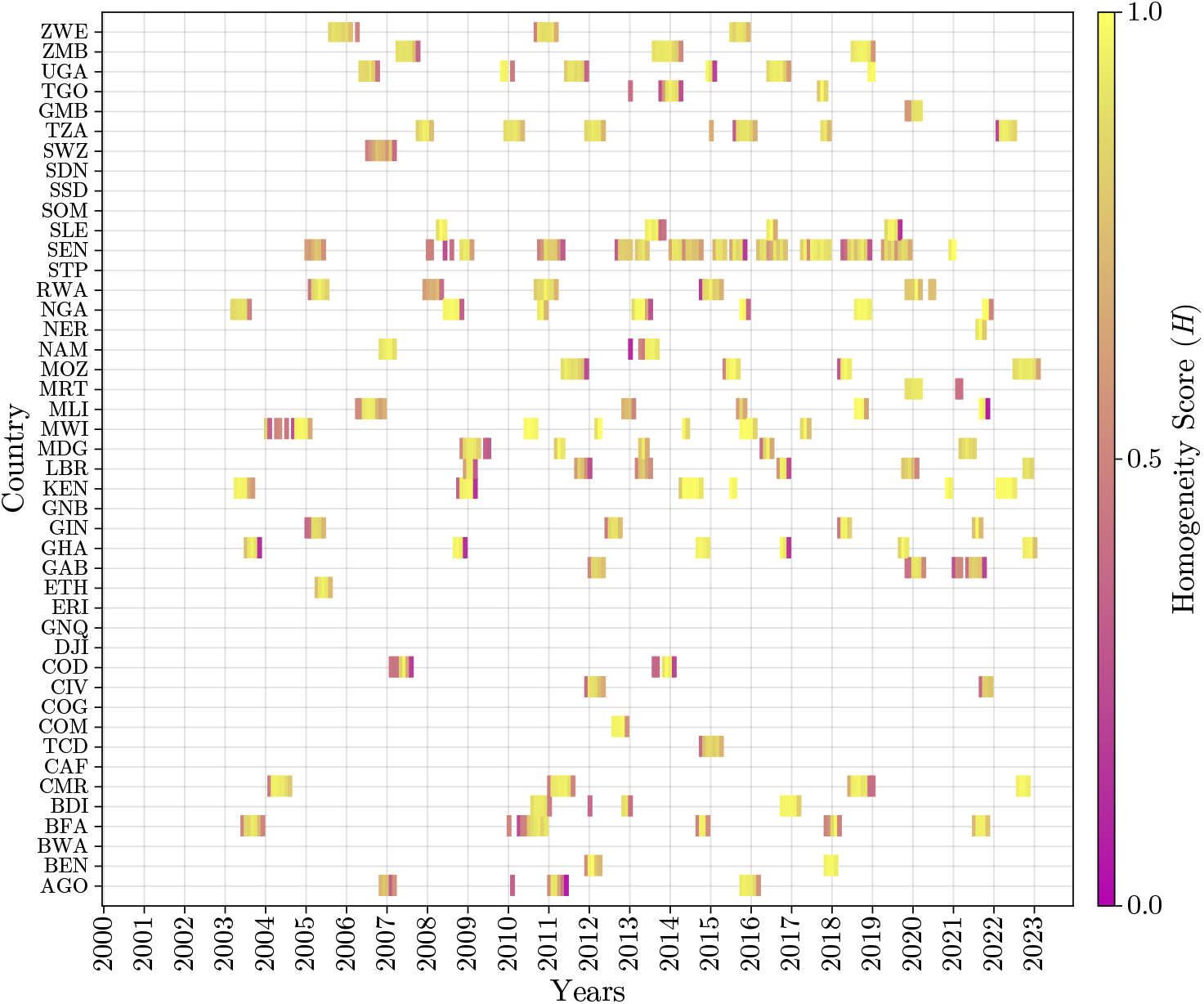
Homogeneity scores for household surveys spanning 44 countries.

Overall, we find that homogeneity scores are high for a majority of countries indicating that temporal homogeneity assumptions hold for most surveys. Lower scores are also observed for periods in the first and last months of a survey period and correspond to cases where the survey period begins close to the end of a given month or ends slightly after the start of the next month. Expectedly, these cases have smaller sample sizes and thus are usually more localised in their spatial distribution. We also find that these results are relatively robust with the selection of resolution *d***x**.

### (c) Observation uncertainty inflation

A benefit of fitting models with a Bayesian approach is that it allows for the inclusion of the level of confidence of observations in the regression. Therefore, a relative weighting of the importance of each observation can be accounted for when performing model calibration. A simple way to achieve this is to assume the errors in observations are normally distributed,

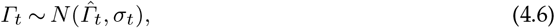

where 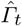 is the SNF model estimate of net crop, *Γ*_*t*_ is the observed survey estimate of net crop, and *σ*_*t*_ describes the level of confidence of the survey observation where larger values of *σ*_*t*_ indicate lower confidence.

Whilst the above approach is easy to implement, choosing a sensible value of *σ*_*t*_ is not obvious. In the context of malaria survey where each observation *Γ*_*t*_ is treated as the weighted mean aggregate of all individual surveyed households in a given month, one a reasonable option would be to set *σ*_*t*_ equal to the standard error,

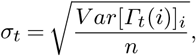

where *V ar*[*x*_*t*_(*i*)] is the variance of the estimated statistic *Γ*_*t*_ at month *t* with respect to *n* households indexed by *i*.

The usage of the standard error appears reasonable at first glance. However, when one accounts for the relatively large sample size of surveys (typically on the order of 10^3^), the estimates for *σ*_*t*_ become extremely small an thus attributes a very high level of confidence to a given observation. In the context of ITNs where most measured values are quantised and metrics of interest tend of be ratios between integers (e.g. nets per person in household), the small value of *σ*_*t*_ may not be a sufficiently conservative estimate of the uncertainty associated with the observation and heavily constrains the model which can lead to overfitting when directly used for regression. This problem is exacerbated in cases where there is volatility in values between observations in consecutive months within the same survey, the cause of which may be multiple. In the absence of sufficient regularisation, this can lead to the risk of overfitting of disaggregation ratios to account for the high level of perceived confidence due to large sample sizes *n*.

To address this issue, we propose two different data pre-processing approaches to more sensibly constraint the model. This first approach consists of applying a moving average filter across observations in consecutive months to reduce the magnitude of fluctuation between months. This approach is simple to implement and strength of the smoothing can be controlled with the width of the smoothing window. However, this simplicity comes at the expense of directly changing the mean of the observation data, which may negatively affect the fit of the model.

A second more nuanced approach is to inflate the uncertainty associated with each observation to account for the temporal mismatch between consecutive observations. In doing so, one avoids directly altering the mean of the observation and thus ensures that the centre of the observation is still preserved. Temporal mismatch between monthly estimates of a given survey can be attributed to two main sources: (1) natural attrition of nets, and (2) sampling errors. Because the former forms a part of the net crop dynamics in the form of net attrition, temporal mismatch between monthly estimates cannot be directly used to define an appropriate inflation factor. Instead, one should calculate an inflation factor accounting for the effect of natural net attrition.

For simplicity, we assume nets decay exponentially with a halflife of some time scale *τ*. Therefore, the number of nets remaining from an initial net crop of *X*_0_ is described by,

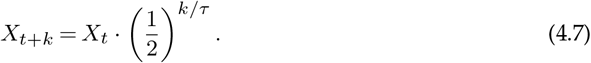

For variance inflation, let *n*_*i*_ be the number of survey observations corresponding to month *i*. Calculate 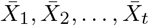 and 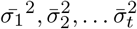, the mean estimates and standard error for *t* separate months based on a single household survey respectively. Assuming the normality of observations within each month, the observed net crop *X*_*i*_ in any given month *i* can be modelled as,

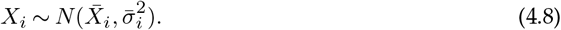

For each month *i*, draw *n*_*i*_ samples 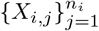 and construct forecast and backcasted estimates based on the assumed attrition curve,

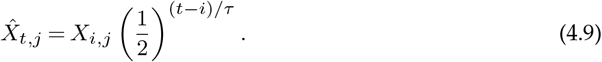

Assuming Equation 4.7 is a sufficiently adequate representation of the attrition time scales, survey estimates with minimal temporal mismatch due to sampling errors should yield small values of 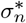 where

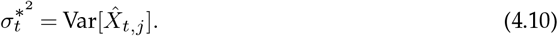

The unadjusted variance 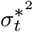 can be calculated for each month to get an estimate of variability excluding the effects of net attrition.

To account for variability due to sample size, the variance 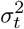 used in the regression is calculated by inflating 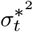 with a factor *η*_*t*_,

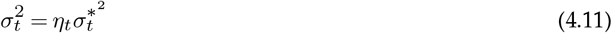

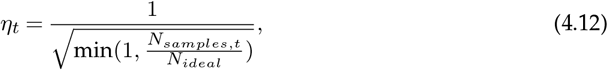

where *N*_*samples,t*_ is the number of samples in the survey month *t*, and *N*_*ideal*_ is a selected threshold to describe the number of samples in any given month that are required to have full confidence in the monthly survey data. For *N*_*samples,t*_ *> N*_*ideal*_, *η*_*t*_ = 1 and the monthly data is deemed to have a sufficiently large sample size and does not require additional variance inflation.

The selection of an exponential decay in the above approach is intentional as the memoryless property allows for convenient calculations of the forward and backward projections in Equation 4.9. We note that a difference in the choice of attrition function for variance inflation compared to those used directly in the SNF is not necessarily problematic as it is only used to inflate the uncertainty. Mismatches between both attrition functions will simply result in more conservative estimates of uncertainty. For all the analyses in this paper, we select an assumed half life of *τ* = 1.5 years, and *N*_*ideal*_ = 4000.

## 5. Model Fitting

### (a) Iterative Expectation-Maximisation (EM) Algorithm

The SNF flow model presents several challenges for model fitting. Firstly, when taken in its entirety with both redistribution and net attrition components, the forward evolution of the SNF model is not easily auto-differentiable. If one wishes to employ a Bayesian approach, apart from explicitly stating and deriving the corresponding likelihood equations, a convenient alternative would be to treat the SNF model as a semi-blackbox. The regression of model parameters can be achieved by utilising a general Markov Chain Monte Carlo (MCMC) method such as the random walk Metropolis-Hasting algorithm. However, direct application is not amenable for the SNF in the proposed formulation due to the large number of model parameters that scales by the length of the analysed time series.

Rather than employing a full Bayesian approach to tackle the high-dimensional regression problem, we tailor the choice of fitting algorithm for each parameter based its relative importance in the model and need for uncertainty quantification. To do this, we separate the parameters in the SNF model into three main classes

- **Direct strong model parameters (***β, ϕ, κ*^(*i*)^, *τ*^(*i*)^): These parameters are presumed to have the largest direct impact on the evolution dynamics of the model and may also meaningful mechanistic interpretations for understanding net dynamics (e.g. net retention time). Therefore, a full Bayesian treatment should be employed to estimate parameter values and associated uncertainty.
- **Direct weak model parameters:** These parameters have direct but relatively short time scale impacts on the model dynamics (e.g. *α* parameters initial conditions, missing net estimates) and whose effects are limited or wane over time. Whilst convergence of these parameters are desirable, uncertainty quantification is beneficial but not necessary
- **Indirect model parameters:** These are parameters that locally affect model dynamics, but are not easily amenable to full uncertainty analysis due to the imposed constraints of the problem. This class consists of the distribution ratios, which must sum to one. They are parameters which provide additional flexibility of the model but do not affect large time scale dynamics and do not strongly benefit from a full uncertainty quantification.

The SNF model is fit using an iterative expectation-maximisation (EM) algorithm. To reduce the dimension of the problem when performing MCMC, the indirect model parameters (distribution ratios 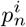) are first fixed to get initial posteriors of direct strong and weak model parameters (*β, ϕ, κ*^(*i*)^, *τ*^(*i*)^, *α* …). Conditioned on these initial estimates, an expectation cost function (E-step) is defined and subsequently optimised (M-step) to obtain updated point estimates of the indirect model parameters 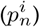. These steps are alternated until convergence.

i. Initialise values for the distribution ratios (indirect model parameters) 𝒟 ^(0)^ = *p*_*t*_ drawn from a random uniform distribution and normalised such that 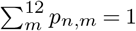.
ii. Estimate the conditional posterior distributions 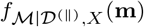 using a Bayesian sampling algorithm (e.g. random walk Metropolis-Hastings).
iii. Draw *N*_*s*_ samples 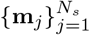 from the posterior *f*_ℳ|𝒟,*X*_.
iv. Define the loss function *L* (E-step) with respect ℳ:

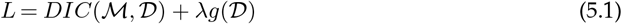

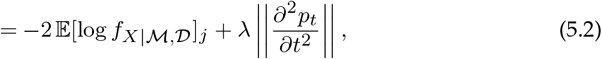

where *DIC* is the posterior log likelihood and *g* is a regularisation term.
v. Numerically calculate the conditional partial derivative 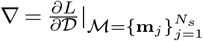 (M-step)
vi. Update estimate for 𝒟 ^(*k*)^ according to a set learning rate *α*_*LR*_.

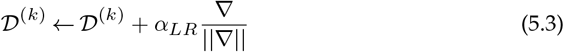
vii. Repeat steps 3-6 for *N*_*step*_ iterations and take final iteration as new estimate for 𝒟 ^(*k*+1)^. This concludes the end of first iteration epoch.
viii. Repeat steps 2-7 for *N*_*epoch*_ iterations or until *L* increases between epochs.

### (b) Convergence and Uniqueness

The complexity of the fitting algorithm and number of parameters presents concerns regarding the convergence behaviour and uniqeuness of the fitted values. This is particularly true for the distribution ratios *p*_*t*_ where point estimates are provided rather than a posterior distribution. In this section, we test the algorithm and demonstrate the numerical stability of its fit when calibrated against a malaria survey dataset. We focus our tests on the modelling of ITN coverage in Nigeria. This country has been chosen due to its demonstrated temporal homogeneity and relatively complete set of observations. The complete dataset for Nigeria consists of annual delivery and distribution data from 2000-2023 and 8 household surveys spanning a total of 39 months.

A total of 20 fits were calculated with randomly initialised monthly disaggregation ratios *p*_*t*_ taken from a uniform distribution and normalised such that Equation 4.2 is met. The algorithm is run up to a maximum of 10 iterations with 30 steps of stochastic gradient descent and 80,000 MCMC joint posterior draws during each iteration. To test the convergence behaviour and uniqueness of solution, we calculate the mean final distribution ratio across all the fits 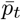 and calculate the RMSE for each *i*^*th*^ model fit 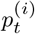 against 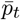,

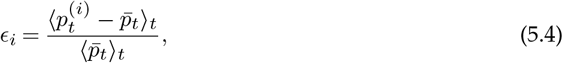

where ⟨… ⟩_*t*_ refers to the time average. The results of the ensemble of fits are shown in Figure 9.

**Figure 9.**
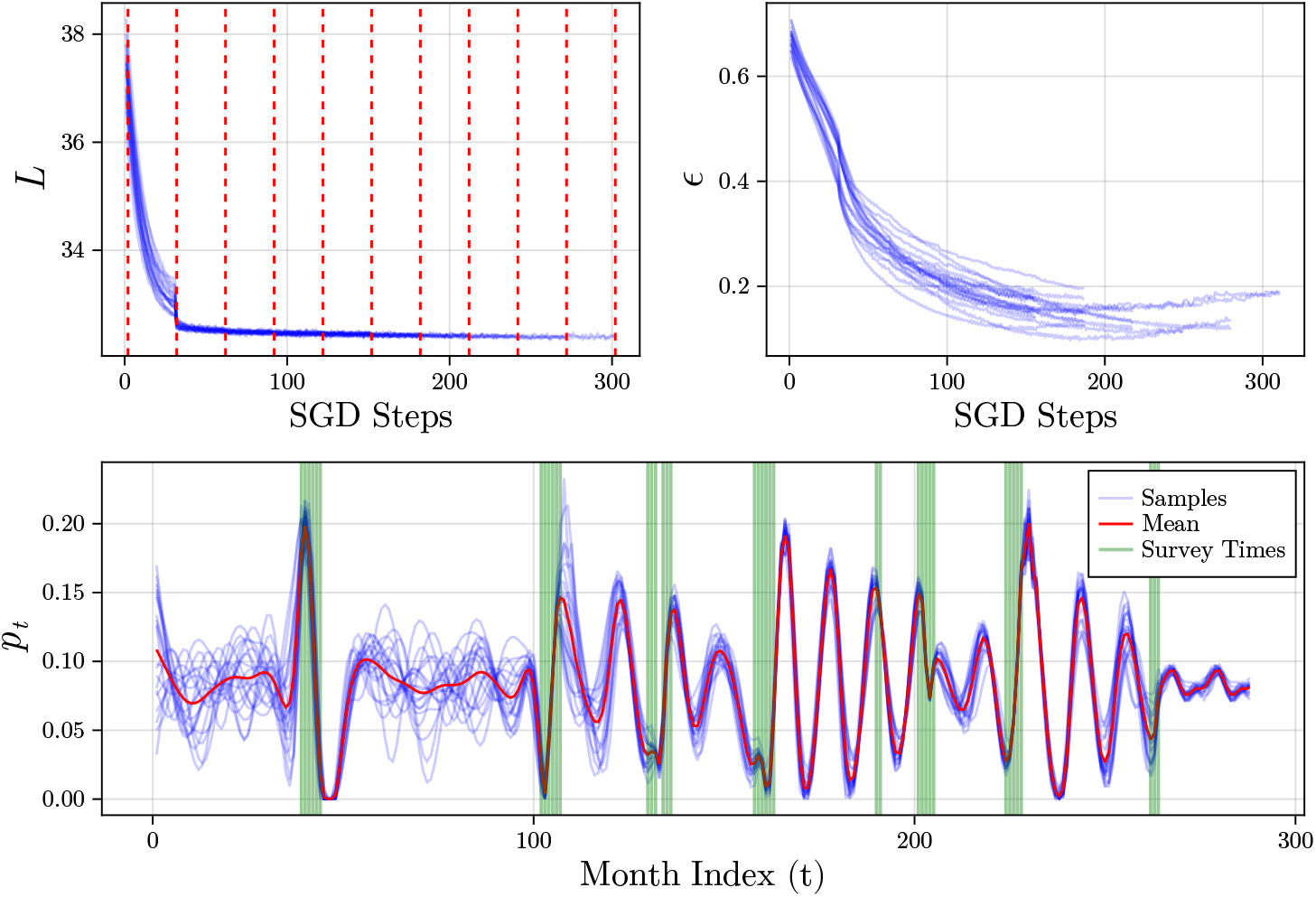
Convergence of loss *L* and *ϵ* with number of SGD steps. Vertical bars represents points where Bayesian resampling of parameters is doe. Collection of 20 final estimates of *p*_*t*_ are shown at the bottom with consistent values near survey data points (green vertical bars). Mean distribution ratio 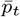 is shown in red.

Overall, the EM algorithm demonstrates very quick convergence with fits reaching within *ϵ* = 0.13 of the final value within 7 iterations (210 SGD steps) with auto-termination occurring as early as 6 iterations. The largest improvement occurs during the first iteration with a discontinuity in the loss curve due to the MCMC resampling of model parameters. Closer inspection of the final distribution ratios also show a consistency between each model fit despite having randomised initial conditions. However, the consistency is only apparent where there is available household survey data to calibrate against. Furthermore, deviations in data sparse locations of the time series do not adversely affect regressed ratios in data rich areas. This suggests that despite its high-dimensionality, the regressed solutions are unique and that distribution ratios only have temporally local effects on SNF dynamics.

### (c) Data Requirements for Model Fit

One concern in the application of SNF models in the ITN context is the risk of overfitting as the number of model parameters tend to far outnumber available of data points used to calibrate the model. In the case for Nigeria, only 39 points are used to calibrate the entire model. Even if strong regularisation is applied, the amount of the data required for calibration of SNF models is not obvious.

To investigate this problem, the full SNF model is trained with varying amounts of calibration data *n*. Three different data omission methods are used: (1) Chronological - increasing amount of data included in chronological order, (2) Reverse Chronological - increasing amounts of data included starting from the most recent, (3) Random - select *n* random observations to include in the training data set. Models are trained on the Nigerian dataset containing two different net types (cITN and LLIN). For the random data omission case, results are aggregated over 10 fits. To measure convergence, we examine the posterior distribution of net half lives of each net type with respect to increasing amounts of training data *n*. Results are given in Figure 10.

**Figure 10.**
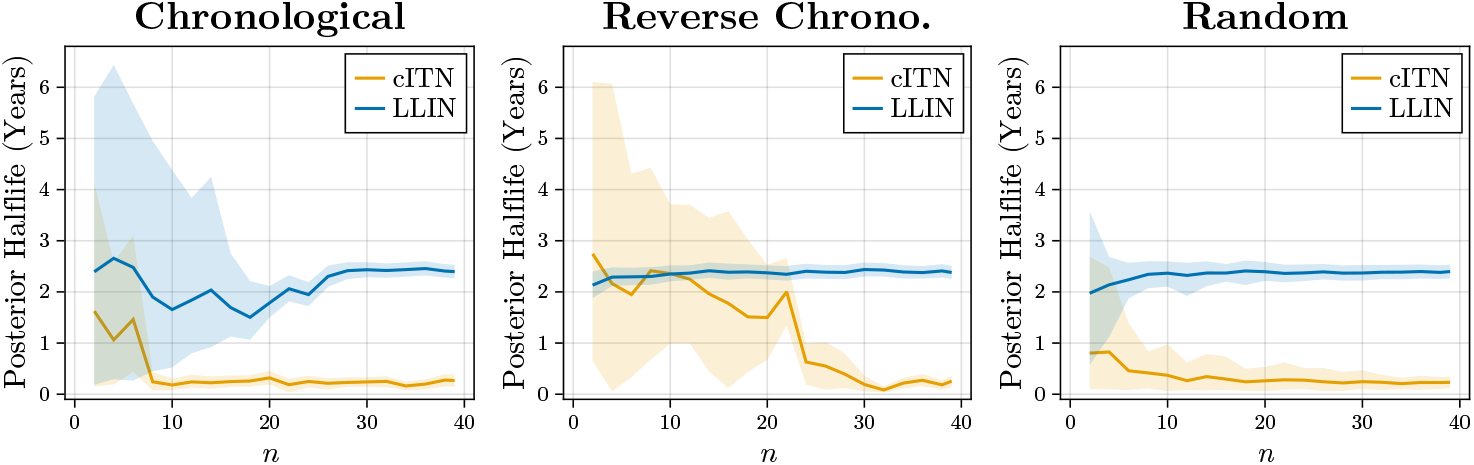
Estimated posterior half life for SNF models trained on increasing amount of observation data points *n*.

In all three test cases, convergence of the mean occurred for relatively modest amounts of training data. In the random case, this was observed in as few as 10 data points, with larger amounts of data required for the chronological and reverse chronological cases. For the former, cITN convergence occurs very quickly whilst LLIN does not converge until *n >* 20. This is due to fact that LLINs did not see widespread adoption in the first half of the dataset (pre-2010) resulting in little to no useful observation data. However, convergence is quickly achieved after the first LLIN observations are included. The opposite is true for the reverse chronological case where instead cITN convergence is delayed until data points far enough back in history containing cITN observations are included. The larger amount of data required for the chronological and reverse chronological compared to the random case is expected as observation data (monthly scale) are temporally clustered based on surveys. The order-agnostic random data inclusion scheme thus allows a larger span of time to be include for calibration without requiring a large amounts of data to be included.

The relatively modest data requirements for SNF model calibration is a welcome feature in the context of ITN modelling where data is typically sparse. One reason for this convergence behaviour can be understood using dynamical systems reduction of the SNF model presented in Section 2(c).

The form given by Equation 2.12 describe how the vector field of the stock and flow dynamics is largely determined by the input driving signal *u*(*t*). Net attrition only act as small perturbations to the vector field. Since *u*(*t*) is fixed and determined based on input distribution data, the dynamics of the stock and flow model is heavily constrained. This implies that a large number of model parameters do not heavily alter the model outputs. Additionally, because the perturbations *ξ* are small, relatively modest amounts of data is required to calibrate model outputs *x* to survey observations.

## 6. Conclusion

Compartmental SNF models are a key component in ITN coverage models. Owing to their mathematical flexibility, the design of these compartmental models have often relied on ad hoc approaches with relatively sparse mathematical justification. Fitting these models is also difficult as they traditionally require the calibration of many model parameters using limited and sparse training data, which may span multiple sampling resolutions. As such, this poses several questions on the choice of mathematical form, convergence behaviour and identifiability of model parameters.

In this paper, we provide a comprehensive analyses of the compartmental “stock-and-flow” (SNF) model commonly employed in ITN coverage models. Using tools from dynamical systems theory and time series analysis, two theoretical reductions of the SNF model are presented and subsequently used to identify governing components in the SNF model. Using a combination of numerical simulations and theoretical arguments, we demonstrate the importance of attrition functions in SNF models and present a case for the use of flexible two parameter functions over simpler one-parameter forms.

In addition to the discussion on attrition functions, we provide a sensible method for disaggregating training data to reconcile temporal resolution mismatch and account for potential mismatches and uncertainty in temporally close observations. To demonstrate its application, an expectation-maximisation algorithm is proposed to fit the model. Our results show that model fits are robust and convergence is achieved for relatively modest levels of training data. Model convergence is briefly discussed from a dynamical systems approach. Modest data requirements are attributed to the dominance of distribution input time series in model outputs with model parameters only representing small perturbations to the underlying vector field to be learned.

## Data Availability

Input data: The household-level survey data used in this analysis is publicly available from the DHS (https://dhsprogram.com/) and MICS (https://mics.unicef.org/) websites. The national-level-aggregated survey data were gleaned from reports available at the MIS website (https://www.malariasurveys.org/. Data on manufacturer delivery of nets are available from the AMP Net Mapping Project (https://allianceformalariaprevention.com/working-groups/net-mapping/). Data on NMCP distribution of nets were provided by WHO.

https://github.com/eugenetkj98/MITN-Public

## Acknowledgements

This work was supported, in whole or in part, by the Bill & Melinda Gates Foundation [INV-055192]. The conclusions and opinions expressed in this work are those of the author(s) alone and shall not be attributed to the Foundation. Under the grant conditions of the Foundation, a Creative Commons Attribution 4.0 License has already been assigned to the Author Accepted Manuscript version that might arise from this submission. Please note works submitted as a preprint have not undergone a peer review process. This work also includes funding support from the Australian Government, National Health and Medical Research Council (Award No: GNT2025280). This activity has also been supported by the Western Australian Future Health Research and Innovation Fund (Grant ID WANMA/Ideas2023-24/9)

